# A Comprehensive Typing System for Information Extraction from Clinical Narratives

**DOI:** 10.1101/19009118

**Authors:** J. Harry Caufield, Yichao Zhou, Yunsheng Bai, David A. Liem, Anders O. Garlid, Kai-Wei Chang, Yizhou Sun, Peipei Ping, Wei Wang

## Abstract

We have developed ACROBAT (Annotation for Case Reports using Open Biomedical Annotation Terms), a typing system for detailed information extraction from clinical text. This resource supports detailed identification and categorization of entities, events, and relations within clinical text documents, including clincal case reports (CCRs) and the free-text components of electronic health records. Using ACROBAT and the text of 200 CCRs, we annotated a wide variety of real-world clinical disease presentations. The resulting dataset, MACCROBAT2018, is a rich collection of annotated clinical language appropriate for training biomedical natural language processing systems.

## 1 Introduction

A diverse set of text documents embodies our expanding knowledge of biological phenomena, including human health and disease. Every type of observation, from the semi-structured results in experimental studies to the detailed narratives in clinical case reports (CCRs) or electronic health records (EHR), is growing in volume, variety, and complexity. Any single human reader must therefore perform extensive labor when using clinical narratives to comprehensively answer biomedical questions, especially when comparing observations across medical subdomains. The structured data yielded by foundational advances in biomedical information extraction (BioIE) is of great assistance in addressing this challenge: they can consistently identify concepts and events within specific domains and tasks. However, interpreting biomedical text with the greatest accuracy depends upon (sub)domain knowledge: e.g., the word “elongated” may describe different types of clinically concerning but surgically correctable deformities, such as an elongated tricuspid valve leaflet in the heart or an elongated styloid process in the skull (i.e., Eagle’s syndrome (1)). Identification of specialized terminology with highly contextual semantics within biological and clinical text remains an open challenge for BioIE methods.

Recognizing the need for resources supporting adaptation of BioIE to clinical narratives, we seek to standardize the entity, event, and relation types within clinical text with a high degree of granularity. Previous work toward addressing this challenge has defined entities though both comprehensive lexicons and high-level types (e.g., the UMLS Semantic Network’s 133 semantic types and 54 relationship types (2), the MeSH (3) controlled vocabulary, or terms in the Medical Dictionary for Regulatory Activities (4)). These resources support thorough biomedical document indexing but face limitations as a basis for clinical IE, particularly as they are intended to represent general biomedical phenomena and observations. More specific clinical coding systems (e.g., LOINC (5), HL7 FHIR (6), ICD-10 (7), or the newly released ICD-11 (8)) exhaustively represent medical events but may require more detail than that afforded by clinical text: a report stating that “microscopic examination of a biopsy revealed a typical LCH” first requires understanding that the LCH is the rare cancer Langerhans-cell histiocytosis, yet accurate assignment of an ICD-10 code demands knowledge of this disease’s progression through the body (i.e., whether it is unifocal or disseminated). Extensive knowledge of each coding system’s intricate hierarchy is therefore necessary for their application to IE in clinical narratives.

We therefore sought to design a typing system for clinical text capable of reflecting the diverse vocabulary and phenomena described within a clinical document without requiring direct connections to curated concepts or terminology. We believe this approach is ideal for designing practical BioIE systems as it primarily reflects the semantics of terminology as it is used rather than an exact correspondence between a set of vocabulary and their expected meaning. A context-driven approach is also intuitive for clinical domain experts. In the above example with the word “elongated”, for example, it is crucial to consistently identify *elongated* as a modifier of a specific anatomical entity in each case, rather than simply the entities as *elongated tricuspid valve leaflet* and *elongated styloid process*. Though the distinction imparted by this level of granularity appears minor, it is not common practice among the few clinical text datasets available for public use.

Our result is a system for Annotation for Case Reports using Open Biomedical Annotation Terms (ACROBAT). ACROBAT is a set of concepts, categories, and relations (in short, a typing system) for representation of medical language. We have specifically designed this system to identify terms and spans within CCRs, though the types are sufficiently broad to afford generalization to other document types written in biomedical language. This system does not correspond to a single ontology, i.e., it does not incorporate a controlled vocabulary or lexicon. Regarding IE methods, we see this system as foundational to implementation of model-driven BioIE, and with this goal in mind we have completed a set of annotations for each of 200 CCRs. To our knowledge, this collection is the only collection of CCRs deeply annotated for both entities and relations. The number of documents in our corpus exceeds that of other deeply-annotated biomedical corpora (e.g., the 97 documents within the CRAFT corpus (9), though we note our documents include only case report text rather than complete manuscript text) while remaining a manageable size for a variety of biomedical IE explorations. All data are available on Figshare (https://doi.org/10.6084/m9.figshare.c.4652765).

## 2 Methods

### 2.1 General design of the ACROBAT clinical typing system

ACROBAT is appropriate for manual annotation, automated labeling (i.e., named entity recognition and relation extraction), or a combination of both. For each document in a corpus, ACROBAT should be used to label all words and phrases in the document corresponding to one or more of the types described in Tables 1 and 3 below. We make a distinction between events and entities: events occur during specific points in time (i.e., they may be arranged into a timeline) while entities are other meaningful text spans, often those modifying or describing properties of events. As a general guideline, the smallest span describing a single entity or event is labeled: for example, in the phrase *massive heart attack*, the labeled event is *heart attack*, as “heart attack” refers to a specific condition and *attack* alone is too general. The term *massive* describes is an entity in its own right; the term is a modifier of the Severity type. Words and phrases are labeled even if they do not specifically discuss a patient, e.g., if the authors discuss hypothetical situations or a patient’s family members. This increases the total number of annotated instances and therefore the total pool of potential training examples. Labels may also overlap where appropriate or when multiple labels apply.

**Table 1:** Event types.

| Type | Description |
| --- | --- |
| Activity | Patient actions and habits. |
| Clinical_event | A clinical activity other than a medical procedure, often involving a change of Nonbiological_location. |
| Diagnostic_procedure | Any procedure done primarily in order to obtain more information. |
| Disease_disorder | Any disease. Essentially a higher-level medical condition potentially including a collection of symptoms. |
| Lab_value | Any result of a laboratory test or a diagnostic result, including any units or values present. |
| Medication | Any pharmaceutical treatment. Used with Administration and Dosage entities. |
| Outcome | The patient’s clinical outcome. |
| Sign_symptom | Any symptom or clinical finding. |
| Therapeutic_procedure | Any procedure done primarily in order to address or alleviate a symptom or disease. This includes surgery, long-term therapies, and supporting procedures (e.g., intubation). |
| *Time expressions* |  |
| Date | A time expression ending at a specific day in time. |
| Duration | A time expression describing a period of time, generally specifying an event has occurred continuously over the given duration. |
| Time | A time expression describing a specific point in time. |
| Other_event | Any event with clinical relevance that does not fit into any of the above categories. |

**Table 2:** Property types.

| Type | Description |
| --- | --- |
| Polarity | Expresses certainty and/or completeness of an event’s occurrence (MAYBE_POS, MAYBE_NEG, UNCERTAIN) or its negation (NEG). |
| Trend | Expresses whether a event has decreased (DEC), increased (INC), or remained the same (STAY) over time, as explicitly stated within the text. |

**Table 3:**
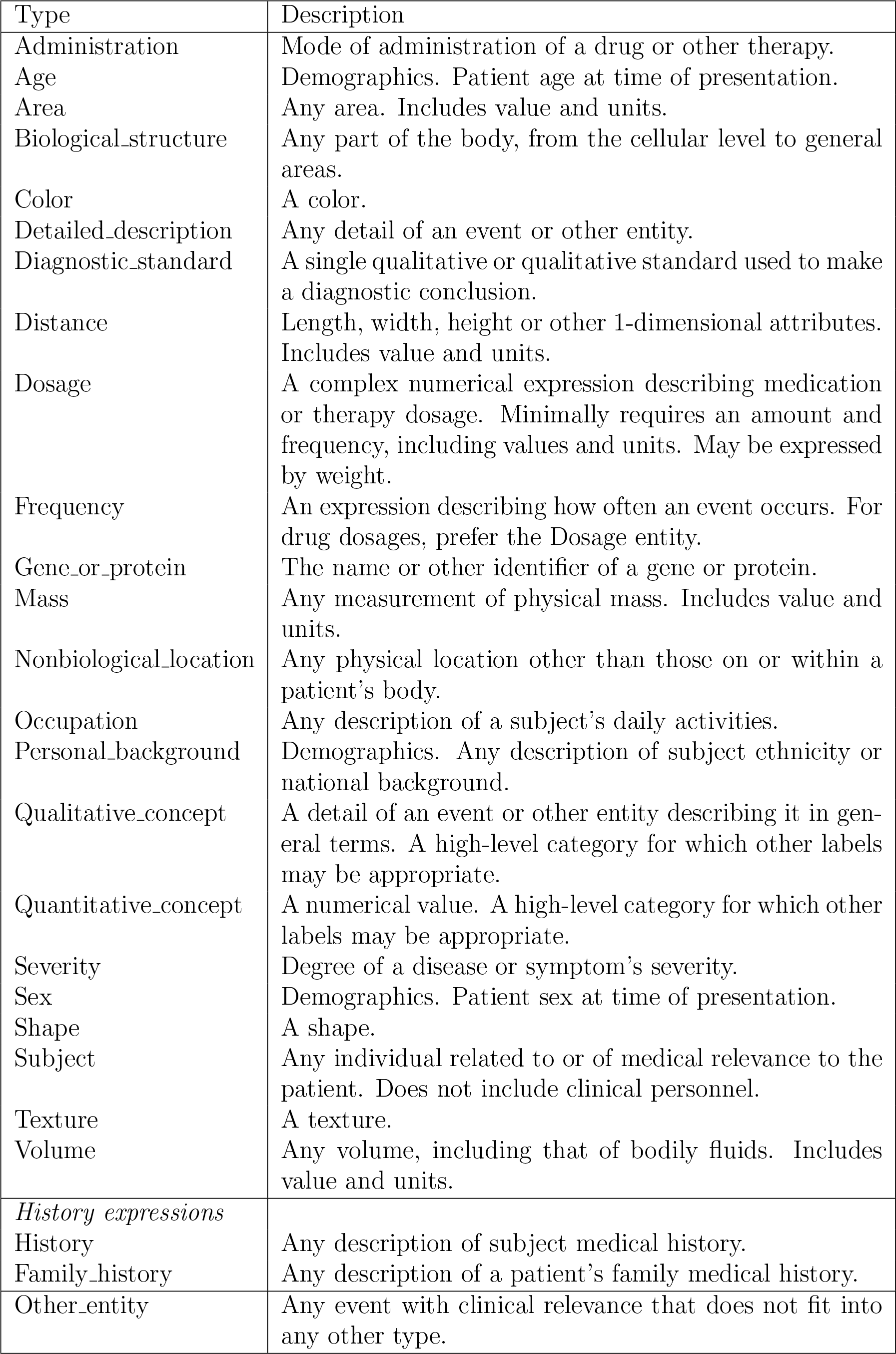
Entity types.

ACROBAT incorporates relations for semantic purposes, coreference resolution, and temporal order (see Table 4). Semantic relations cover all instances in which an entity modifies or results from another event or entity in any manner. For coreferences, an event/entity and any of its coreferences within a single document are linked through pairwise relations. Temporal order defines events within a continuous time series (e.g., *event 1* → *event 2* → *event 3*). Directionality is meaningful in ACROBAT and all relations (with the exception of Identity relations) are directed. ACROBAT also supports event properties for indication of changes over time or event negation (see Table 2). In cases where an abbreviation is present (e.g., *optical coherence tomography (OCT)*), the full name (*optical coherence tomography* and the abbreviation (*OCT*) are labeled as separate events and connected with an Identity relation.

**Table 4:**
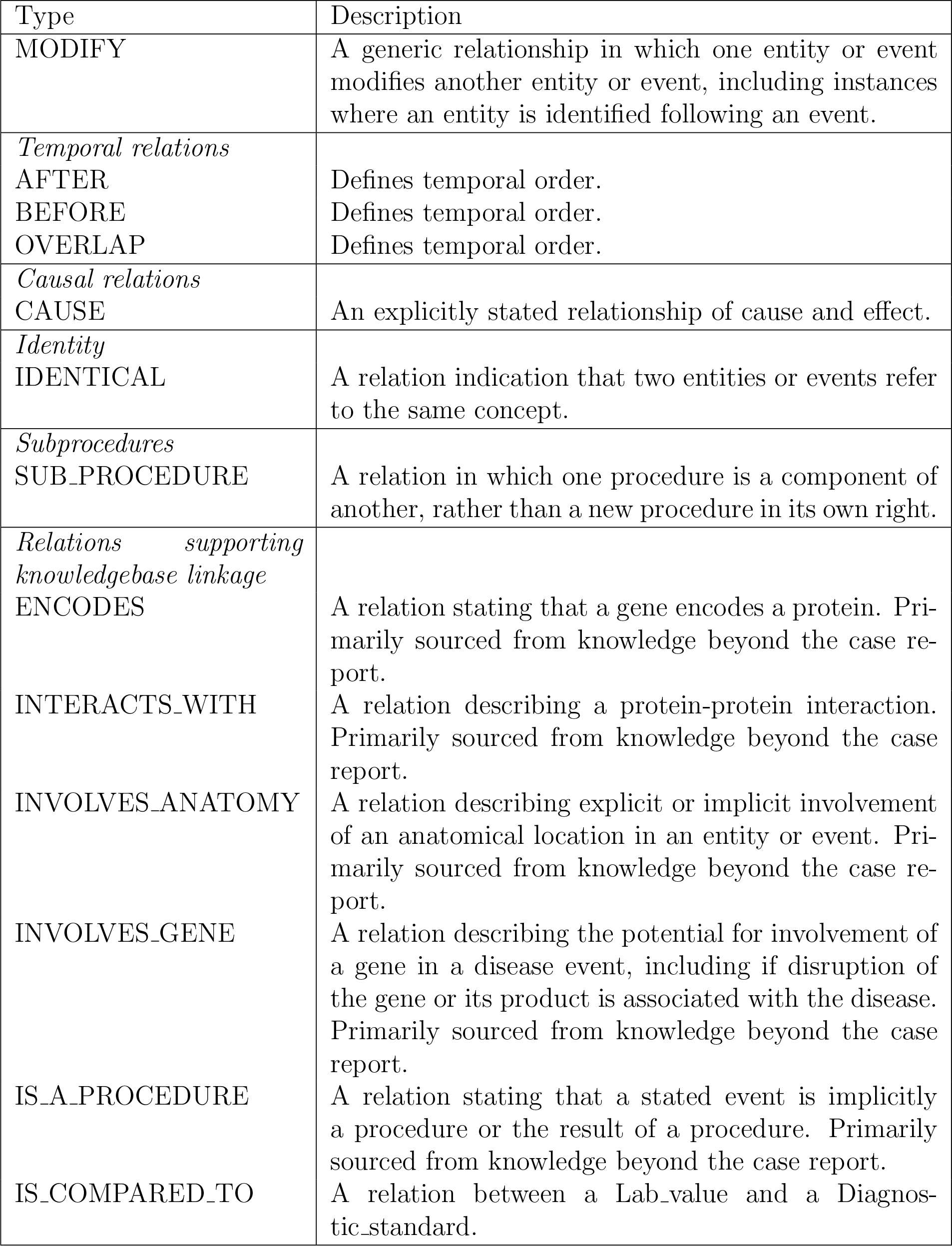
Relation types.

### 2.2 Events in the clinical typing system

Events (Table 1) include words or phrases indicating a discrete activity or occurrence in a document. In this section, we describe each event category, delineate annotation rules, and provide examples. See Table 1 for summaries of event types and Figure 1 for an example of event annotation.

**Figure 1:**
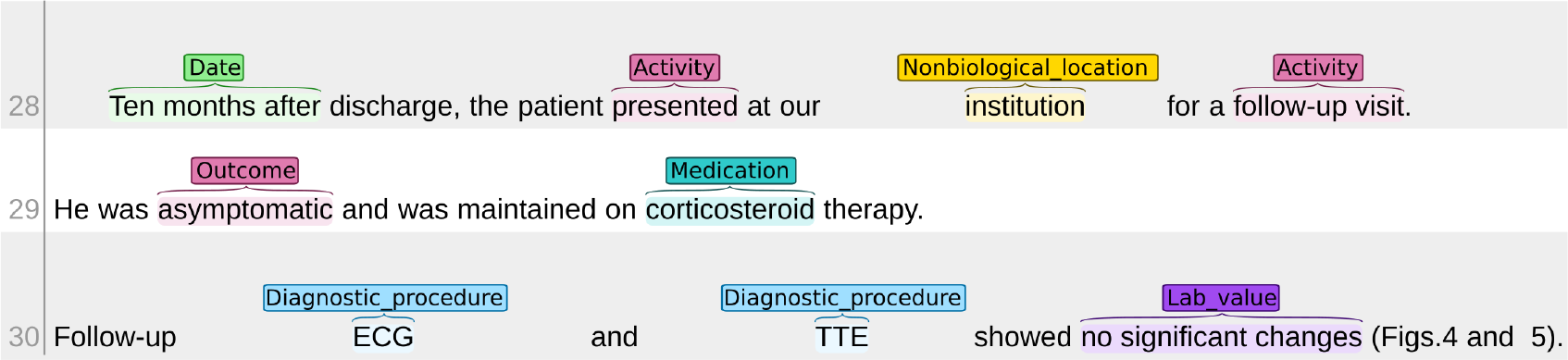
Example visualization of event annotation in a clinical case report. The source report describes a case of Still’s disease, a rare autoinflammatory condition (10). All annotations shown here are events, with the exception of *institution*, which is a Nonbiological_location. Labels are colored by type.

#### 2.2.1 Activity

These events include any word or phrase describing a physical activity performed by a subject once, multiple times, or regularly (e.g., a habit), outside an explicitly clinical context. These activities may or may not refer to accidents, e.g., the phrase *fell out of bed*. Additional examples include *dancing, falling from a bicycle, smoking, drinking*, or *eating*.

#### 2.2.2 Clinical_event

These events include any word or phrase describing clinical events relating to administrative procedures and practices outside a diagnostic or therapeutic context. A change in Nonbiological_location is often involved or implied. Examples include *presented, transferred, admitted*, or *discharged*.

#### 2.2.3 Diagnostic_procedure

These events include any word or phrase comprising the name of any procedure performed for diagnostic purposes, i.e., to collect more information about a patient or any symptom. The definition of a diagnostic procedure in this scheme is intentionally broad in order to capture numerous terminology variations and implicit tests: it includes formal names of procedures, physical examinations, imaging, lab tests, and diagnostic screens for specific conditions. For example, in the phrase “C-reactive protein was not elevated”, *C-reactive protein* is a diagnostic procedure due to context despite the lack of an explicitly stated test. Procedures may yield qualitative and/or quantitative results, expressed as Lab_value events connected to the Diagnostic_procedure through a MODIFY relation. Subprocedures are defined using the SUB_PROCEDURE relation type. Additional examples include *clinical examination, colonoscopy, iron concentration, ultrasonography*, or *electrocardiogram*.

#### 2.2.4 Disease_disorder

These events include any word or phrase comprising the name of a disease, health disorder, or injury. These include specific names (e.g., *type 2 diabetes*), high-level conditions (e.g., *infection*; *metastasis*), categories of disease (e.g., *mitochondrial disease*), infective agents (e.g., *Salmonella enterica*), anatomical abnormalities (e.g., *stenosis*), and specific varieties of neoplastic growth (e.g., *adenocarcinoma*). In general, Disease_disorder events describe conditions which may involve a collection of signs or symptoms. Additional examples include *myocardial infarction, hydrocephalus, autonomic dysregulation, speech deficits*, or *perforation*.

#### 2.2.5 Lab_value

These events include any word of phrase comprising, in any format, any result of a diagnostic procedure, e.g., a blood pressure of *130/100 mmHg*, ejection fraction of *40%, low* platelet count, or *slowly regained* renal function. Lab_value events may also relate to signs and symptoms, e.g., *5* tumors, tachycardic to *170 bpm*, or *enlarging* mass. Additional examples include: histological examination was *normal*, body mass of *60 kg, positive* for adipophilin, or cancer (*T3N1M0*).

#### 2.2.6 Medication

These events include all names of medications and pharmaceutical treatments, including general classes of compounds and drug therapies. Examples include *medication, chemotherapy, vancomycin, Tris-hydroxymethyl aminomethane*, or *sedation*.

#### 2.2.7 Outcome

These events include any brief but explicit explanation of a case outcome. This is contextual, as any description of a patient’s state at the beginning of a disease progression is by definition not an outcome. Outcome events are therefore located near or at the end of documents. Clinical cases may frequently omit patient outcomes, particularly if they are not known or are indicated by other events (e.g., an autopsy, though not labeled as an outcome, implicitly states a patient has died). Examples include *asymptomatic, excellent result, death*, or *expired*.

#### 2.2.8 Sign_symptom

These events include any description of a symptom or clinical finding. These words or phrases may include general conditions whether they are localized or not (e.g., *pain, fever, rash*, or *dizziness*); descriptions of abnormal or unexpected conditions, potentially accompanied by specific diagnostic procedures and Lab_value events (e.g., *anemia, weight loss, tachycardia*, or *leukocytosis*); localized conditions dependent upon anatomical context (e.g., extremities were *warm, stenosis* of the left main coronary artery, or *enlarged* gallbladder); or uncharacterized abnormal growths (e.g., *mass, lesions*, or *tumor*). Results of diagnostic procedures, histological examinations, and imaging describing clinically concerning findings are also Sign_symptom events, e.g., *foam cells, contrast defect* was observed, or *abnormality*. Additional examples include *unable to move, erythematous* mucosa, *gastrointestinal symptoms*, or *metastases* in the right adrenal gland.

#### 2.2.9 Therapeutic_procedure

These events include any word or phrase comprising the name of any procedure performed for therapeutic treatment purposes, i.e., to address the cause or symptoms of a disease or disorder. These procedures may support other procedures (e.g., *intubation*), including surgery (e.g., *opened, drainage tube*, or *cleaned*). Additional examples include *radiation therapy, arterial embolization, surgical repair, anastomosis*, or *blood transfusions*.

#### 2.2.10 Time expressions

Date events are time expressions including any word or phrase denoting a specific day, whether in absolute or relative time (i.e., specifying an exact date or a day with a date contingent on knowing other dates or times, respectively). These are similar to Duration expressions in practice but have discrete endpoints and may describe events that did not occur continuously during the specified time (e.g., *A* happened, then *B* happened on this date). Examples include *April 7 1982, Monday*, or *two months later*.

Duration events are time expressions including any word or phrase denoting a period of time. They generally indicate a seperate event has occured continuously or regularly over the course of the time period. Examples include *one week, two months*, or *overnight*.

Time events include any work or phrase describing a specific point in time at granularity more specific than days. Examples include *3 PM, 15 minutes later, 40 hours later*, or *a few minutes later*.

#### 2.2.11 Other_event

Other events of clinical note occur regularly among labeled events. These events are labeled when they relate to specific patients (e.g., do not describe general phenomena across patient populations or medical practice), extenuating circumstances, and/or when their meaning cannot be determined from the surrounding text. Examples include *national shortage, difficulty* in operating surgical equipment, *resistance*, or *earthquake*.

### 2.3 Properties in the clinical typing system

Properties (Table 2) are attributes of a single event. All events may include values for neither, one, or both properties.

The Polarity property indicates negation or how certain a document’s author’s are regarding the reported event. All events implicitly have a Polarity of `POS` by default, indicating no uncertainty; a value of `NEG` indicates an event was specified but did not happen (e.g., the patient reported no *pain*; *surgery* was not performed). Polarity values of `MAYBE_POS`, `MAYBE_NEG`, or `UNCERTAIN` indicate an event may have, may not have, or has little certainty to have completely occurred, respectively (e.g., *influenza* was suspected but not confirmed; almost complete resolution of bilateral *infiltrates*, suspicion of *embolism*).

The Trend property indicates change in an event over time. Its value is one of `DEC`, `INC`, or `STAY`, representing any stated decrease, increase, or constancy, respectively. Examples include: *symptoms* subsided over time, progressive *dysphagia*, or *fever* was sustained.

### 2.4 Entities in the clinical typing system

Entities include words or phrases that do not completely constitute a clinical event on their own but generally modify an event or subject. See Figure 2 for an example of their usage along with event annotations.

**Figure 2:**
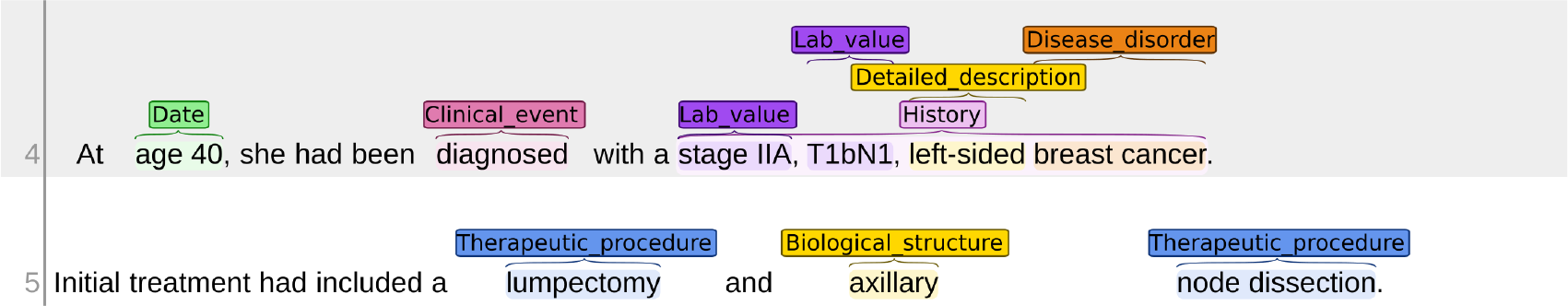
Example visualization of entity annotation in a clinical case report. The source is a clinical case report describing a case of breast cancer (and, later in the report, cardiomyopathy) (11). Relations are omitted and labels are colored by type.

#### 2.4.1 Physical description modifiers

These entities include any word or phrase serving as a modifier of an event’s physical properties. Where applicable, entities include both values and units. Multiple sets of units and values may be present. Values may be expressed quantitatively or qualitatively. Physical description modifier entities include the following types:

- Area: any expression of two-dimensional space or surface area. Examples include *10 cm*^*2*^, *2 cm x 7 cm*, or *2*.*5 cm x 2*.*4 cm in diameter*.
- Color: examples include *red, salmon-colored, pigmented*, or *yellowish*.
- Distance: any expression of one-dimensional length. Examples include *3*.*6 cm in height, up to 15 mm in diameter*, or *50 mm long*.
- Mass: examples include *50 mg* or *ten kg*.
- Shape: examples include *ovoid, spindle-shaped, crystalline*, or *bubbly*.
- Texture: examples include *soft, smooth*, or *ground-glass*.
- Volume: any expression of three-dimensional space. Examples include *50 ml, 2*.*8 cm x 2*.*8 cm x 2*.*5 cm*, or *600 cc*.

#### 2.4.2 Procedure modifiers

These entities modify Medication events or relevant procedures (e.g., radiation therapy). They have the types Administration and Dosage. Administration includes any expression indicating how a material or procedure is administered to a patient, including method or mode of entry; examples include *injection, intravenous, mouth rinse, parenteral*, or *p*.*o*. (i.e., by mouth). Dosage includes any expression indicating amount and/or frequency of a material’s administration. Please note that, in absence of context, these values may appear identical to Mass or Volume entities; examples include *500 mg daily for 3 days, 60 UI/kg/day, 5000 cGY in 25 fractions, high dose, 4 boluses/month, four infusions*, or *800 mg*.

#### 2.4.3 Patient modifiers

These entities modify the patient. For this reason, they do not generally participate in relationships, but are instead implicitly understood to modify the document’s primary subject. These entities include:

- Age: examples include *65-year-old, 20 years old, newborn*, or *teenage*.
- Family_history: any set of words or phrases describing medical history and events experienced by someone related to the patient. Note that events within these spans are also annotated as necessary, e.g., the span *heart failure* within *his brother died of heart failure* is a Disease_disorder event. Examples include *family history of hypertension, her aunt was a carrier of the mutation, his parents and two older brothers were healthy*, or *parents were cousins*.
- History: any set of words or phrases describing the patient’s medical history. As with Family_history, events within the span receive their own annotations. Examples include *pregnancy, diabetes, no history of smoking, past medical history was unremarkable*, or *history of controlled hypertension*.
- Occupation: any expression describing the patient’s daily activities or surrounding environment. Examples include *police officer, student, office worker, works on a farm*, or *worked in a chemical plant for 30 years*.
- Personal_background: any expression describing the patient’s national, cultural, or ethnic background. Examples include *Japanese, Caucasian, from the UAE, American, Spanish descent*, or *African-American*.
- Sex: any expression describing the patient’s biological sex at the time of the report. This may be implicit in documents describing generally sex-specific diseases such as obstetric conditions, e.g., *primipara* is a valid annotation for Sex as it implicitly states the patient is female. Additional examples include *woman, male*, or *boy*.

#### 2.4.4 Biological_structure

Entities of this type include any words or phrases describing an anatomical location of any size. Examples include *blood vessel, left femoral vein, liver, left side of body, mitochondria, subcutaneous*, or *mediastinal lymph nodes*.

#### 2.4.5 Detailed_description

These entities include any modifier not included in other entity types. They often include phrases differentiating Sign_symptom, Disease_disorder, or procedure events: they may describe conditions (e.g., *sudden* cardiopulmonary arrest), extent (e.g., *right-sided* paralysis), subtype (e.g., *group B* streptococcus; *pleomorphic* carcinoma), or type of diagnostic (e.g., *serum* uric acid). Additional examples include *emergent* intubation, *fiberoptic* bronchoscopy, *spontaneous* mesenchimal haematoma, *congenital* bleeding disorder, or *recent* travel.

#### 2.4.6 Frequency

Entities of this type describe how often an event occurs. Examples include *twice a week, daily, occasionally*, or *intermittent*.

#### 2.4.7 Nonbiological_location

These entities include any description of a location in physical, geographic, non-anatomical terms. These terms include names of clinical practitioners (e.g., *cardiologist*) as these statements generally refer to a specific location as well. Examples include *our hospital, cardiac intensive care unit, Boston, home*, or *emergency department*.

#### 2.4.8 Severity

Entities of this type include any description of a symptom or disease’s degree of severity. Examples include *severe, mild, slight, extensive*, or *profuse*.

#### 2.4.9 Subject

These entities identify individuals in the document other than the patient. They may include family members. They generally do not include clinical personnel. Examples include *his uncle, mother*, or *her cousin*.

#### 2.4.10 Other_entity and general modifiers

Entities of clinical relevance but not appropriate for other types are labeled with either Qualitative_concept or Quantitative_concept if they modify another event or entity. All others receive Other_entity annotations.

#### 2.4.11 Entities supporting knowledgebase linkage

Two entity types, Diagnostic_standard and Gene_or_protein, support connections to biomedical knowledge bases. Neither is used in clinical report annotation as they are rarely stated explicitly and therefore constitute domain knowledge. Diagnostic_standard entities include any standard to which the result of a diagnostic procedure (i.e., a Lab_value) is compared. They may be expressed in natural language, e.g., *normal lactate concentration is less than 1*.*6 mMol/L*. We note that diagnostic standards are often contextual and subject to patient background, including sex, ethnic background, and presence of comorbidities. Linkage to diagnostic standards is therefore intended to serve as a means of additional interpretative context rather than a means of diagnosis. For Gene_or_protein, any identifier of a human gene, protein, or non-coding transcript constitutes an entity. Examples include *BRCA2, P51587* (a UniprotKB identifier), or *PHF8*.

### 2.5 Relations in the clinical typing system

Relations are connections between entities or events. There are two general categories of relations: those used to express the temporal order of events (BEFORE, AFTER, and OVERLAP), and those used to define more specific relationships. Nearly all relationships, unless otherwise specified, are defined through the MODIFY relation. Examples include lumbar MODIFY→ hernias, moderate MODIFY→ hypokinesis, clinical examination ←MODIFY tachypnea, or respiratory rate ←MODIFY 16 breaths per minute. Note that relation directionality is relevant, and in cases where a procedure yields another event, the source of the relation is the result (e.g., the Lab_value *16 breaths per minute*) while the origin (e.g., the Diagnostic_procedure *respiratory rate*) is the target. Several relation types and their contexts are illustrated in Figure 3.

**Figure 3:**
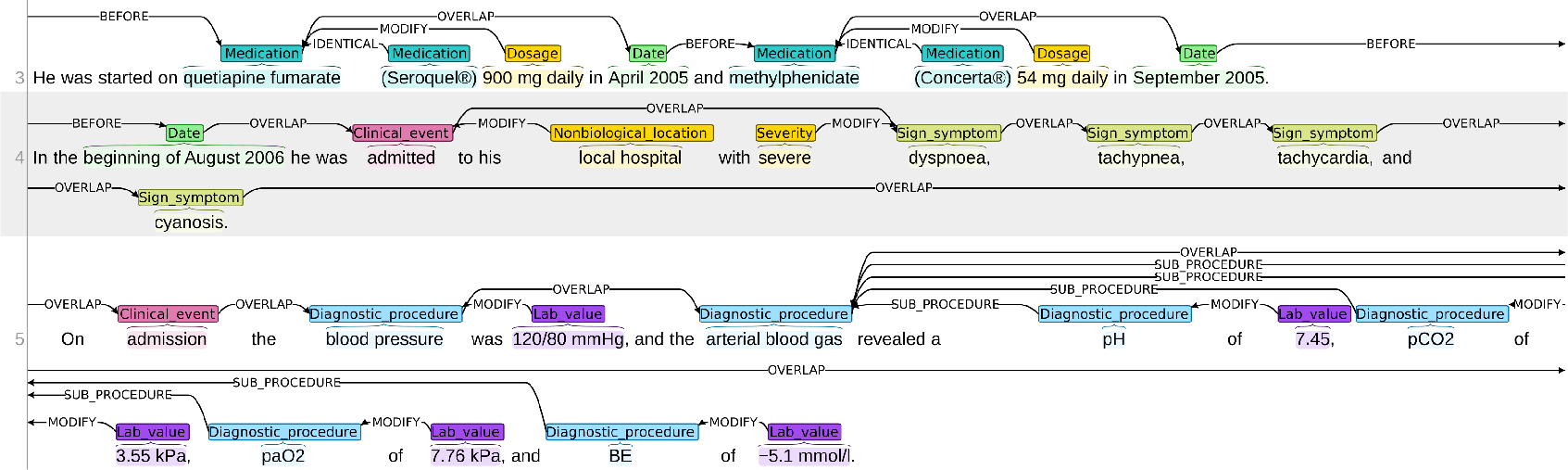
Example visualization of relation annotation in a clinical case report. The source is a clinical case report describing a case of acute dilated cardiomyopathy (12). Temporal, identity, and modifier relations are all included, as are subprocedures (i.e., the *arterial blood gas* procedure has four subprocedures mentioned in the document: *pH, pCO2, paO2*, and *BE*).

#### 2.5.1 Temporal relations

All events are connected by three temporal order relations: BEFORE, OVERLAP, and AFTER. Events happening within the same general timeframe are connected by OVER-LAP relations. A new time frame begins if and when the text describes an event as being before or after another, if a time period of more than 24 hours is indicated, or the patient is moved somewhere (e.g., transferred to a different department, generally noted as a new Nonbiological_location). These changes are annotated with BEFORE or AFTER relations. For example, in the case of “A happens. B happens. C happens.” the connecting temporal relations are (A BEFORE→ B) and (B BEFORE→ C); there is no (A BEFORE→ C) relation included. Time is assumed to progress continuously across sentences. The direction of arrows for temporal order relations is always forward, i.e. from a previous entity to a later one.

#### 2.5.2 Causal relations

Causality is annotated only when expressed explicitly in the text. Examples include *death* ←CAUSE *exsanguination* or *pericardial effusion* CAUSE→ *cardiac tamponade*.

#### 2.5.3 Identity

Words or phrases referring to identical concepts or events are connected with the IDENTICAL relation type. These include coreferences (e.g., *tissue samples* (…) IDENTICAL→ *the samples*) and instances where an acronym follows an event or entity (e.g., *left eye* IDENTICAL→ *LE*).

#### 2.5.4 Subprocedures

Therapeutic or diagnostic procedures performed as part of a more general procedure are connected with the SUB_PROCEDURE relation type. Examples include:

*blood test* ←SUB_PROCEDURE *white blood cell count* or

*laboratory evaluations* ←SUB_PROCEDURE *hemoglobin*.

#### 2.5.5 Relations supporting knowledgebase linkage

The following relation types support connections to biomedical knowledge bases. They are not used in clinical report text annotation.

ENCODES: this relation type supports links between genes and proteins, where both source and target are Gene_or_protein entities, e.g.

*HSD17B10* ENCODES→ *3-hydroxyacyl-CoA dehydrogenase type-2*.

INTERACTS_WITH: this relation type supports representation of protein-protein interactions, e.g. *3-hydroxyacyl-CoA dehydrogenase type-2* INTERACTS_WITH→ *Eukaryotic initiation factor 4A-III*. Both source and target are Gene or protein entities.

INVOLVES_ANATOMY: this relation type represents involvement of a Sign_symptom or Disease_disorder event in an anatomic context, i.e., a connection to a Biological_structure entity beyond those described in the document. Examples include: *pulmonary hypertension* INVOLVES_ANATOMY→ *pulmonary artery* or *pyloric stenosis* INVOLVES_ANATOMY→ *pylorus*.

INVOLVES_GENE: this relation type represents conceptual association between a Disease_disorder and a Gene_or_protein, as defined through domain knowledge. Examples include *hereditary hemorrhagic telangiectasia* INVOLVES_GENE→ *ENG* or *cerebral cavernous malformations-1* INVOLVES_GENE→ *CCM1*.

IS_A_PROCEDURE: this relation type represents a connection between an implied procedure and an explicitly defined one. The source event may be a Diagnostic_procedure or a Sign_symptom implying a procedure was performed, e.g., *lactates* IS_A_PROCEDURE→ *blood lactate concentration*.

IS_COMPARED_TO: this relation type supports connections between Diagnostic_standard entities and Lab_value events, e.g. *140 bpm* IS_COMPARED_TO→ *normal adult resting heart rate is 60 to 100 bpm*.

### 2.6 Coreferences

Rather than denoting specific events, Coreferences label words or phrases referring to previously defined events or entities (i.e., linguistic anaphora). Annotating a coreference therefore defines a relation but takes the form of an event in our system to accomodate labeling of the corresponding text spans. Coreferences must be linked to the last instance of their corresponding referenced term (i.e., their antecedent) with an IDENTICAL relation. Examples include *it, they, the samples*, such that the terms may refer to *carcinoma, the patient’s parents*, or *biopsy results*, respectively. Coreference tags are also used in instances where an equivalent term is used instead of that used previously (e.g., if a *carcinoma* is referred to as a *tumor* two sentences later, *tumor* is labeled as a Coreference) and instances where an identical phrase is repeated but refers to the same event rather than a new occurrence.

### 2.7 Annotation of clinical case reports

In order to prepare a deeply annotated resource of clinical text, we sought to annotate clinical case reports using the schema described above. This work follows from creation of our Metadata Acquired from Clinical Case Reports, or MACCR, set (13): each source document in the new set corresponds to a single entry, and therefore a collection of higher-level metadata, in our MACCR dataset. We therefore refer to our set of deeply annotated CCRs as MACCROBAT2018. For each of 200 documents, we obtained open-access text from PubMed Central, limiting the text portion to that comprising the clinical case (i.e., we did not include any other sections including introduction, discussion, figure/table legends, or supplementary materials). The document count was chosen based on manageability; subsequent releases will add more annotated documents. Each document in the set is named based on the respective PubMed identifier of their source document. All documents were selected based on the following criteria:

- They are present in the MACCR set.
- They concern only a single, human patient each (or, a single mother and child).
- They are in English.

All documents were annotated by at least one of six annotators, all either senior undergraduate university students or post-doctoral researchers. All annotators had previous experience reading biomedical and clinical language. Annotations were checked for format and type consistency upon completion. Annotation was performed through an implementation of the *brat* tool (14); all annotation visualizations presented in this manuscript were created with *brat*. An example of annotation of a CCR is provided in Figure 4.

**Figure 4:**
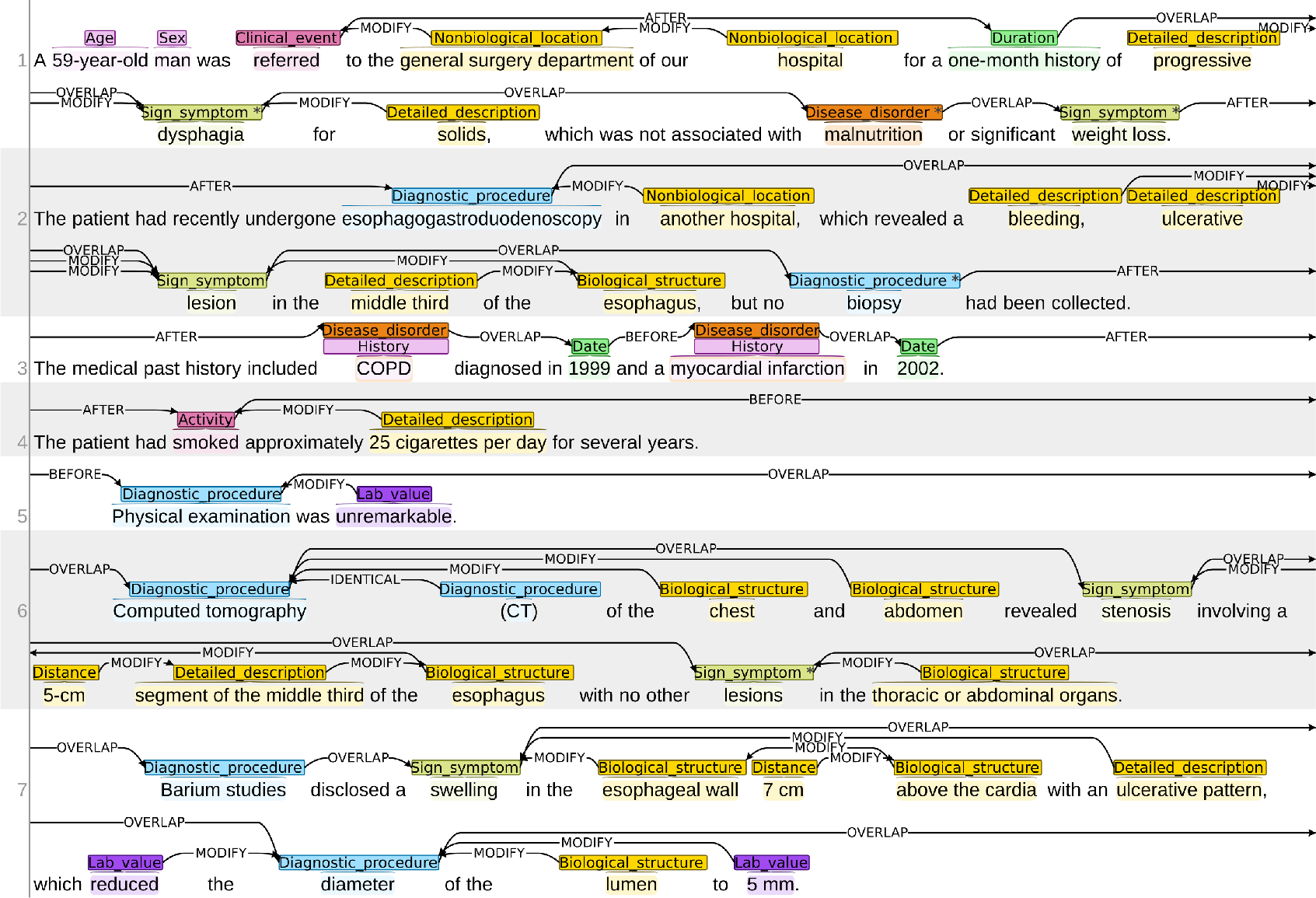
Example visualization of a selection of an ACROBAT annotated document. The source is a clinical case report describing a case of esophageal cancer (15). Labels are colored by type. Labels with asterisks indicate those with property values, e.g., *malnutrition* in the first line has a POLARITY of NEG as this event is mentioned as not having been observed.

The MACCROBAT2018 set can be downloaded at the following location through Figshare: https://doi.org/10.6084/m9.figshare.c.4652765

## 3 Results

The MACCROBAT2018 is intended to serve multiple purposes. This collection of annotations serves as both a demonstration of the ACROBAT scheme: each document is annotated with all appropriate event and entity types as well as relations, thereby providing numerous contextual examples of the typing scheme’s implementation. The set is also ideal for training/testing BioIE methods as it covers a variety of disease presentations and corresponding vocabulary: the 200 documents in the set contain an average of 22.7 sentences, or 4,541 sentences in total, with sentences containing an average of 21.6 single-word tokens and 98,038 tokens in total. Because the text is annotated with multiple label and relation types, it may be used for the initial training of joint models (e.g., a tagger for both diagnostic procedure events and their results).

The MACCROBAT2018 set contains a total of 3,652 sentences and 59,164 annotations of any kind, including event/entity labels and relations. Further counts of annotations by type are detailed in Table 5 for events/entities and Table 6 for relations. Out of all categories and all 200 CCRs, Diagnostic_procedure events occur most frequently, with an average of more than 45 occurrences per document. The set also includes more than 6,700 annotations of signs/symptoms and more than 3,500 lab values.

**Table 5:** Event and entity annotation counts.

| Type | Count |
| --- | --- |
| Activity | 216 |
| Administration | 175 |
| Age | 207 |
| Area | 43 |
| Biological_structure | 2,931 |
| Clinical_event | 1,252 |
| Color | 53 |
| Coreference | 626 |
| Date | 1,462 |
| Detailed_description | 2,901 |
| Diagnostic_procedure | 9,135 |
| Disease_disorder | 2,724 |
| Distance | 122 |
| Dosage | 362 |
| Duration | 560 |
| Family_history | 81 |
| Frequency | 77 |
| History | 392 |
| Lab_value | 3,565 |
| Mass | 2 |
| Medication | 2,152 |
| Nonbiological_location | 354 |
| Occupation | 13 |
| Other_entity | 20 |
| Other_event | 44 |
| Outcome | 84 |
| Personal_background | 57 |
| Qualitative_concept | 41 |
| Quantitative_concept | 31 |
| Severity | 369 |
| Sex | 191 |
| Shape | 65 |
| Sign_symptom | 6,718 |
| Subject | 54 |
| Texture | 46 |
| Therapeutic_procedure | 2,010 |
| Time | 114 |
| Volume | 33 |

**Table 6:**
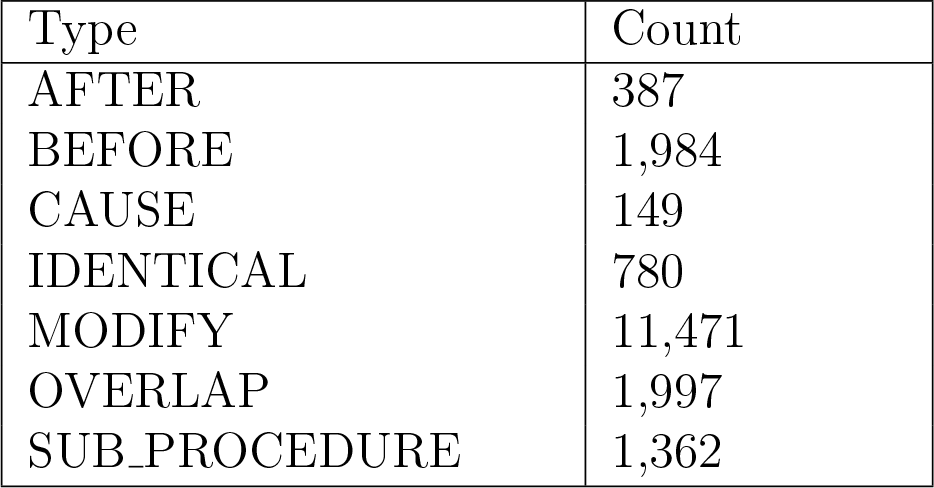
Relation annotation counts.

As compared to other annotated corpora in biomedicine, MACCROBAT includes far more entity and relation types, as well as explicitly defined types for integration with knowledgebases. Sets with similar or greater numbers of coreferences (e.g., the GENIA (16), 2011 i2b2 Coreference Challenge (17), ODIE (18), or CRAFT (9) corpora) have been completed but do not exclusively focus on clinical language and, in some cases, are freely available in their entirety. As coreferences are not the primary focus of our annotations, it may be appropriate to use our set along with other corpora for applications in training coreference resolution models.

## 4 Discussion

A consistent set of concept types is a valuable resource for biomedical informatics in both philosophy and practice. We see the diverse event, entity, property, and relationship types defined by ACROBAT as a way to formalize the specific details particular to a clinical case report or clinical narrative, such that the text within these documents can be treated as structured data. Our goal in designing this system is to provide a means to enforce structure on a variety of concept types within clinical text while not mandating mapping of entities to a particular vocabulary (e.g., terms may or may not be present in SNOMED CT or MeSH) or coding system (e.g., concepts may or may not correspond to ICD-10/11 codes). Additionally, events and entity types may vary in frequency of occurrence in a given text corpus, as is seen with our annotated MACCROBAT2018 resource. We hold that these realities reflect the nature of biomedical language: it is far more variable in its semantics between documents, subjects, and authors than any single index, typing system, or ontology can capture. Therefore, we see ACROBAT and MACCROBAT2018 as a way to manually or computationally enforce structure upon biomedical language, and in doing so, produce resources for training and developing systems for better understanding the concepts within clinical documents and publications.

## Data Availability

All data are available on Figshare as indicated in the manuscript.

https://doi.org/10.6084/m9.figshare.c.4652765

## 5 Acknowledgements

The authors would like to thank Sanjana Murali for insightful input and feedback during the design of ACROBAT, as well as help with annotation. We are also grateful for annotation assistance from Jessica Lee, Linda Do, and Jonathan Quach. No competing interests are declared.

